# Antiviral Antibody Fc Dysfunction Is Linked to Increased IgG1 Fucosylation in Long COVID

**DOI:** 10.64898/2026.09.28.26364044

**Authors:** Rahul Ukey, Ioannis E. Michailidis, Patricia Greenberg, Tracy Andrews, Sung Yun Jung, Haiyan Zheng, William Honnen, Abraham Pinter, Lawrence C. Kleinman, Natalie Bruiners, Maria Laura Gennaro

**Author notes:** Corresponding author: Maria Laura Gennaro, International Center for Public Health, Room W250W, 225 Warren Street, Newark, NJ 07103.

## Abstract

**Background:** Persistent SARS-CoV-2 antigen is a leading hypothesis for post-acute sequelae of SARS-CoV-2 infection (PASC), commonly known as Long COVID. Impaired Fc-mediated antiviral antibody function could contribute to inefficient clearance of infected cells and viral antigen despite preserved antibody levels. Therefore, we tested whether PASC is associated with altered Fcγ receptor-mediated antibody function and differences in IgG1 Fc fucosylation, a major regulator of Fc effector function.

**Methods:** Serum samples from RECOVER-Adult participants with prior SARS-CoV-2 infection (99 PASC, 139 non-PASC) were analyzed for anti-SARS-CoV-2 spike receptor-binding domain (RBD) IgG and subclasses, FcγRIIIa- and FcγRIIa-mediated reporter activity as surrogate measures of antibody-dependent cellular cytotoxicity (ADCC) and antibody-dependent cellular phagocytosis (ADCP), anti-RBD IgG1 Fc glycosylation, and circulating fucose-modifying enzymes.

**Results:** Despite higher anti-RBD IgG and IgG1 abundance, PASC sera showed lower ADCC- and ADCP-associated FcγR reporter activity, including after normalization for antibody abundance. Anti-RBD IgG1 showed greater fucosylation in PASC samples (30 PASC, 34 non-PASC). Serum FUCA2 was lower in PASC samples (24 PASC, 34 non-PASC), whereas FUCA1 and soluble CD16a levels did not differ between the two groups.

**Conclusion:** PASC is associated with impaired antiviral IgG Fc function, increased IgG1 Fc fucosylation, and reduced circulating FUCA2, identifying a glycoimmune axis that may contribute to impaired Fc-dependent antigen clearance and represents a potentially targetable pathway in Long COVID.

## Introduction

Post-acute sequelae of SARS-CoV-2 infection (PASC), commonly referred to as Long COVID, comprises persistent or recurrent symptoms following acute infection and can involve multiple organ systems, with substantial effects on functional status and quality of life^1, 2^. Despite intensive investigation, the mechanisms that sustain PASC remain incompletely defined, and validated mechanistic biomarkers and targeted therapies are still lacking^3^.

Persistence of SARS-CoV-2 antigen or viral reservoirs is a leading mechanistic model for PASC^4, 5^. Antibodies contribute to viral clearance not only through neutralization but also through Fc gamma receptor (FcγR)-dependent effector functions. Engagement of FcγRs on innate immune cells triggers antibody-dependent cellular cytotoxicity (ADCC) and antibody-dependent cellular phagocytosis (ADCP), pathways that promote clearance of infected cells and immune complexes^6, 7^. Thus, qualitative defects in Fc-mediated antibody function could permit antigen persistence even when antiviral antibody concentrations are preserved or increased. Whether Fc-mediated antiviral antibody function is altered in PASC remains understudied.

We tested whether PASC is associated with a qualitative defect in anti-SARS-CoV-2 antibody function and altered IgG1 Fc structure. Using serum from vaccinated adults with prior SARS-CoV-2 infection in the NIH RECOVER-Adult cohort, we first determined whether anti-RBD antibody abundance and FcγR-mediated activity differed between participants with and without PASC and then examined Fc glycosylation and circulating fucose-modifying enzymes as potential molecular determinants of altered antibody function.

## Results

### Study cohort

The study included 238 adults from the RECOVER-Adult observational cohort with prior SARS-CoV-2 infection and COVID-19 vaccination, including 99 participants with PASC and 139 without PASC (non-PASC). The two groups did not differ significantly in age or race/ethnicity, whereas the proportion of women was significantly higher in the PASC group, consistent with the known higher prevalence of Long COVID among women ^8^(**Table 1**). Immunity-related comorbidities, including immunocompromising, autoimmune, and rheumatic conditions, were also more frequent in participants with PASC (**Table 1**). The interval between the most recent SARS-CoV-2 infection and sample collection did not differ between groups, whereas the interval since the most recent COVID-19 vaccination was modestly longer in the PASC group than in the non-PASC group ((231.5 ± 128 days vs. 194.5 ± 105.5 days; *p* = 0.03) (**Table 1**). These differences were considered in subsequent multivariable analyses.

**Table 1.**
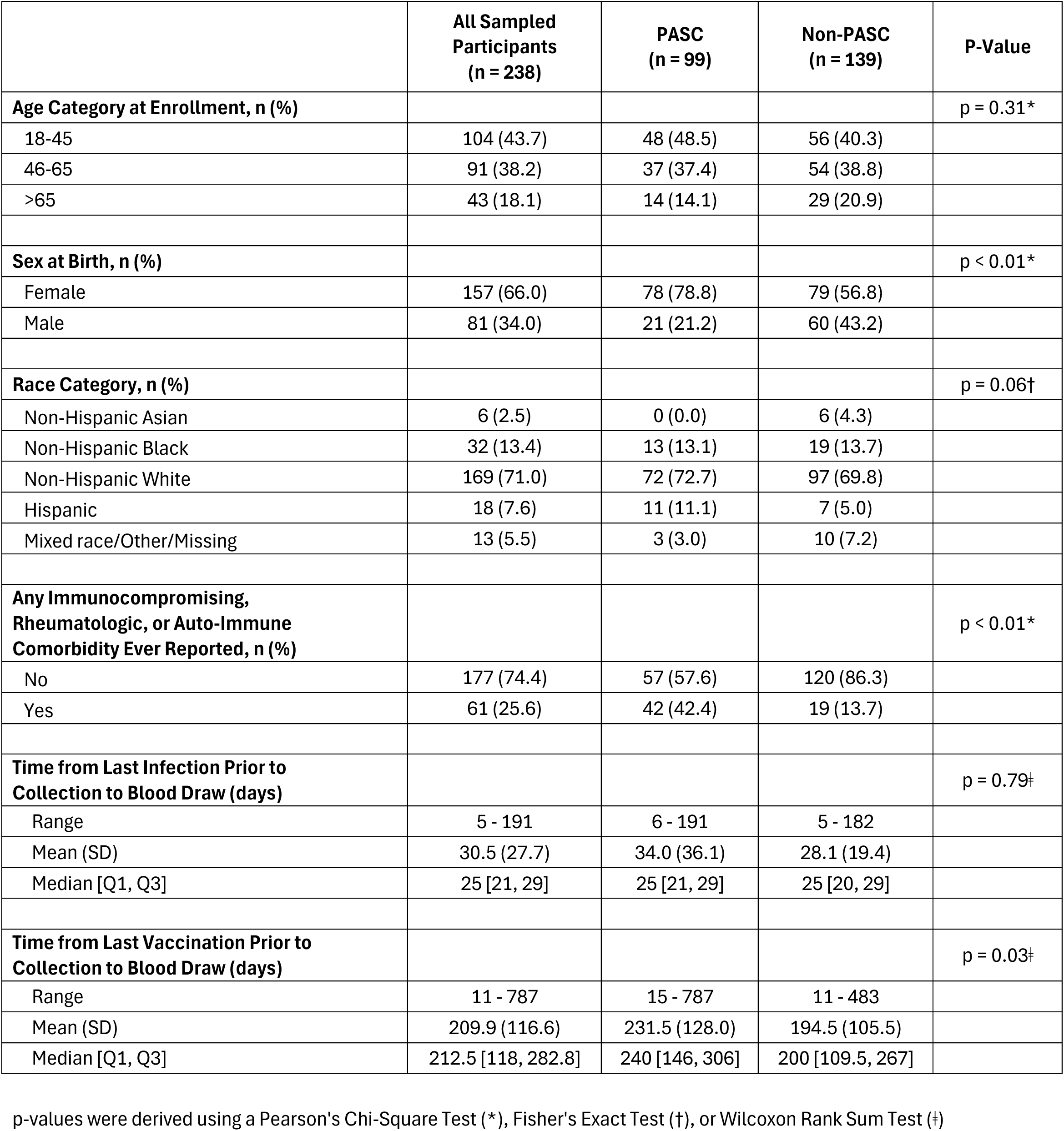
Demographic and clinical characteristics of subjects enrolled in the study, by PASC status.

### Anti-RBD IgG abundance is increased in PASC

We analyzed serum from 99 participants with PASC and 139 without PASC. Anti-RBD IgG levels were higher in the PASC group (**Figure 1A**). Anti-RBD IgG1, the predominant subclass, was also increased in PASC (**Figure 1B**), as was IgG3, whereas IgG2 and IgG4 did not differ significantly between groups (**Table 2**). These findings indicate that PASC was not associated with a quantitative deficiency of anti-RBD IgG.

**Figure 1.**
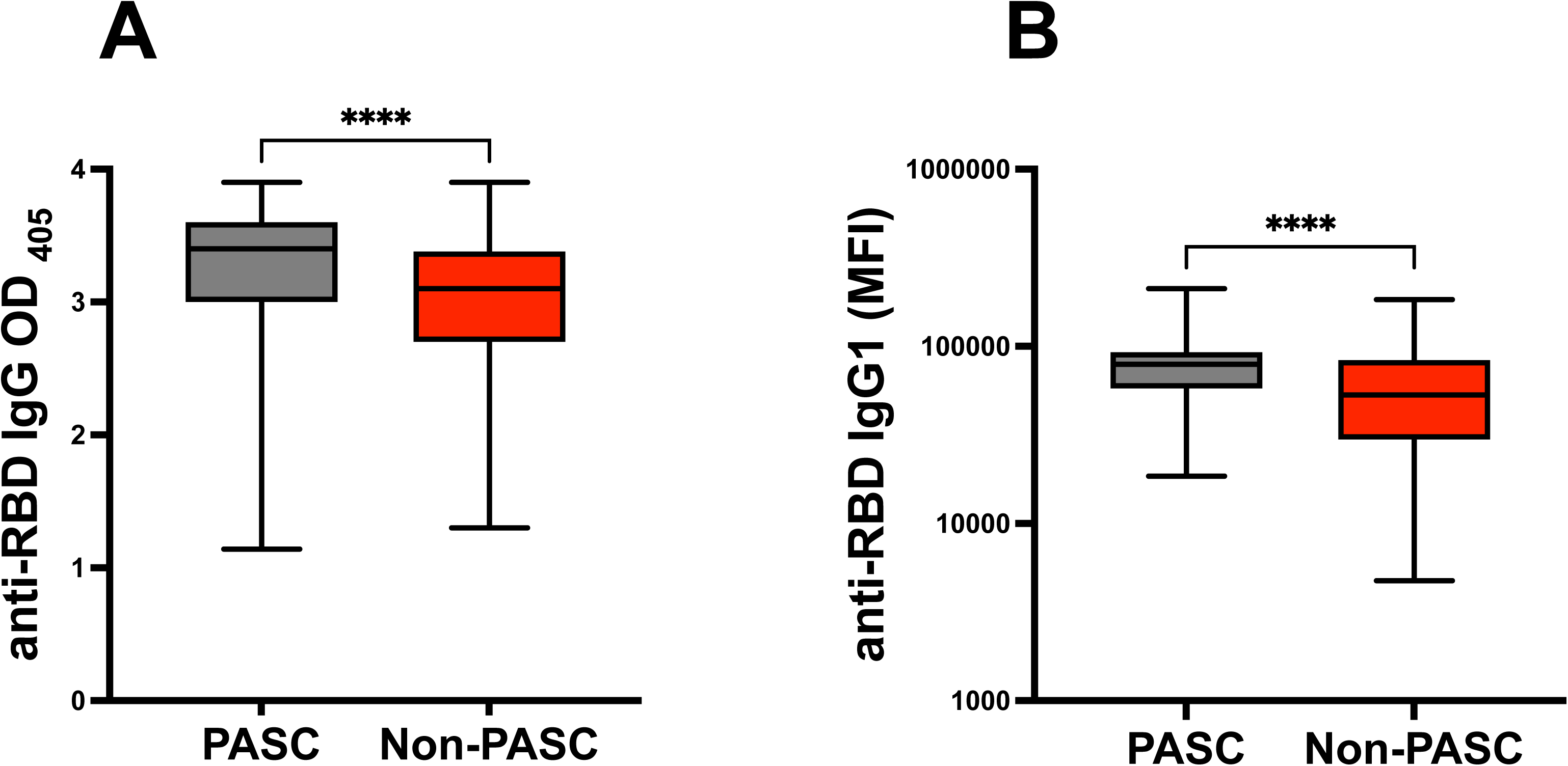
Anti-RBD IgG and IgG1 levels are elevated in PASC relative to non-PASC subjects. Serum was collected from adult RECOVER cohort participants with confirmed SARS-CoV-2 infection and COVID-19 vaccination, classified as PASC (n = 99) or non-PASC (n = 139). (A) Total anti-RBD IgG and (B) anti-RBD IgG1 levels were measured by ELISA/Luminex-based assay (see Methods). Box-and-whisker plots display the median (horizontal line), interquartile range (box, 25th–75th percentile), and full data range (whiskers). Group differences were assessed by the Mann-Whitney U test. *p < 0.05.

**Table 2.** Laboratory outcome measures by PASC status group.

|  | All Sampled Participants<br>(n = 238) | PASC<br>(n = 99) | Non-PASC<br>(n = 139) | P-Value* |
| --- | --- | --- | --- | --- |
| **Anti-RBD IgG** |  |  |  | p < 0.01 |
| Range | 1.1 - 3.9 | 1.1 - 3.9 | 1.3 - 3.9 |  |
| Mean (SD) | 3.1 (0.6) | 3.2 (0.6) | 3.0 (0.5) |  |
| Median [Q1, Q3] | 3.2 [2.8, 3.5] | 3.4 [3.0, 3.6] | 3.1 [2.7, 3.4] |  |
| **ADCC** |  |  |  | p < 0.01 |
| Range | 46.0 - 755.5 | 46.0 - 755.5 | 46.0 - 634.0 |  |
| Mean (SD) | 274.9 (143.3) | 238.7 (135.1) | 300.7 (143.9) |  |
| Median [Q1, Q3] | 270.5 [195.0, 319.0] | 243.0 [133.2, 302.5] | 283.0 [239.5, 338.0] |  |
| **ADCC/Anti-RBD IgG** |  |  |  | p < 0.01 |
| Range | 14.0 - 318.7 | 14.0 - 245.5 | 14.8 - 318.7 |  |
| Mean (SD) | 89.4 (47.0) | 73.3 (40.8) | 100.9 (47.8) |  |
| Median [Q1, Q3] | 81.9 [62.1, 110.4] | 69.4 [49.0, 84.8] | 93.3 [74.2, 130.0] |  |
| **ADCP** |  |  |  | p < 0.01 |
| Range | 41.0 - 1739.5 | 41.0 - 1739.5 | 111.5 - 1563.5 |  |
| Mean (SD) | 763.0 (349.4) | 699.4 (371.6) | 808.2 (326.6) |  |
| Median [Q1, Q3] | 731.5 [524.2, 891.8] | 681.0 [517.0, 815.0] | 795.0 [568.0, 949.5] |  |
| **ADCP/Anti-RBD IgG** |  |  |  | p < 0.01 |
| Range | 15.8 - 835.4 | 15.8 - 596.4 | 81.1 - 835.4 |  |
| Mean (SD) | 245.8 (108.6) | 211.6 (108.1) | 270.1 (102.6) |  |
| Median [Q1, Q3] | 228.0 [174.2, 298.8] | 196.5 [153.4, 233.8] | 263.9 [199.7, 335.4] |  |
| **IgG1** |  |  |  | p < 0.01 |
| Range | 4750.0 - 211939.0 | 18528.0 - 211939.0 | 4750.0 - 183826.0 |  |
| Mean (SD) | 69051.7 (38920.5) | 81902.0 (35824.8) | 59899.4 (38565.1) |  |
| Median [Q1, Q3] | 67227.0 [38268.8, 88161.5] | 79131.0 [58241.5, 92496.0] | 53106.0 [30029.5, 83437.0] |  |
| **IgG2** |  |  |  | p = 0.19 |
| Range | 1668.0 - 780737.0 | 3176.0 - 396920.0 | 1668.0 - 780737.0 |  |
| Mean (SD) | 57129.7 (81996.8) | 57891.9 (69172.5) | 56586.9 (90267.3) |  |
| Median [Q1, Q3] | 24762.0 [9462.5, 78654.5] | 26798.0 [11214.0, 82431.5] | 19280.0 [8296.5, 75238.5] |  |
| **IgG3** |  |  |  | p < 0.01 |
| Range | 1688.0 - 146385.0 | 2887.0 - 146385.0 | 1688.0 - 122837.0 |  |
| Mean (SD) | 12154.6 (16125.2) | 16041.9 (18460.2) | 9385.9 (13638.1) |  |
| Median [Q1, Q3] | 7258.0 [3968.0, 13741.0] | 10750.0 [5471.5, 18856.5] | 5578.0 [3374.5, 10698.5] |  |
| IgG4 |  |  |  | p = 0.57 |
| Range | 1149.0 - 1884930.0 | 1149.0 - 1002070.0 | 1660.0 - 1884930.0 |  |
| Mean (SD) | 332587.8 (384812.3) | 314155.1 (294402.5) | 345716.1 (438571.0) |  |
| Median [Q1, Q3] | 248052.6 [26475.2, 413787.5] | 276570.0 [22438.0, 523623.5] | 239619.0 [36135.0, 388337.0] |  |

\*All p-values were resulted using a Wilcoxon Rank Sum Test

### FcγR-mediated antibody function is reduced in PASC

We next assessed the ability of RBD-specific serum IgG to engage FcγRIIIa and FcγRIIa using standardized Jurkat-Lucia reporter cells as a surrogate measure of ADCC- and ADCP-associated Fc signaling, respectively. Surrogate ADCC and ADCP activities were significantly lower in PASC than in non-PASC sera (**Figure 2, AB; Table 2**). These differences persisted after normalization to anti-RBD IgG abundance and were similarly observed when reporter activity was normalized to anti-RBD IgG1 abundance (**Figure 2, CD; Table 2**). ADCC- and ADCP-associated reporter activities increased with anti-RBD IgG abundance in both groups; however, across the range of anti-RBD IgG levels, PASC samples consistently exhibited lower FcγRIIIa- and FcγRIIa-dependent signaling than non-PASC samples (**Figure 3 AB**). Comparable results were obtained with IgG1 abundance (**Figure S1**). These findings indicate that the reduced FcγR-mediated antibody activity in PASC cannot be explained by differences in antiviral antibody abundance.

**Figure 2.**
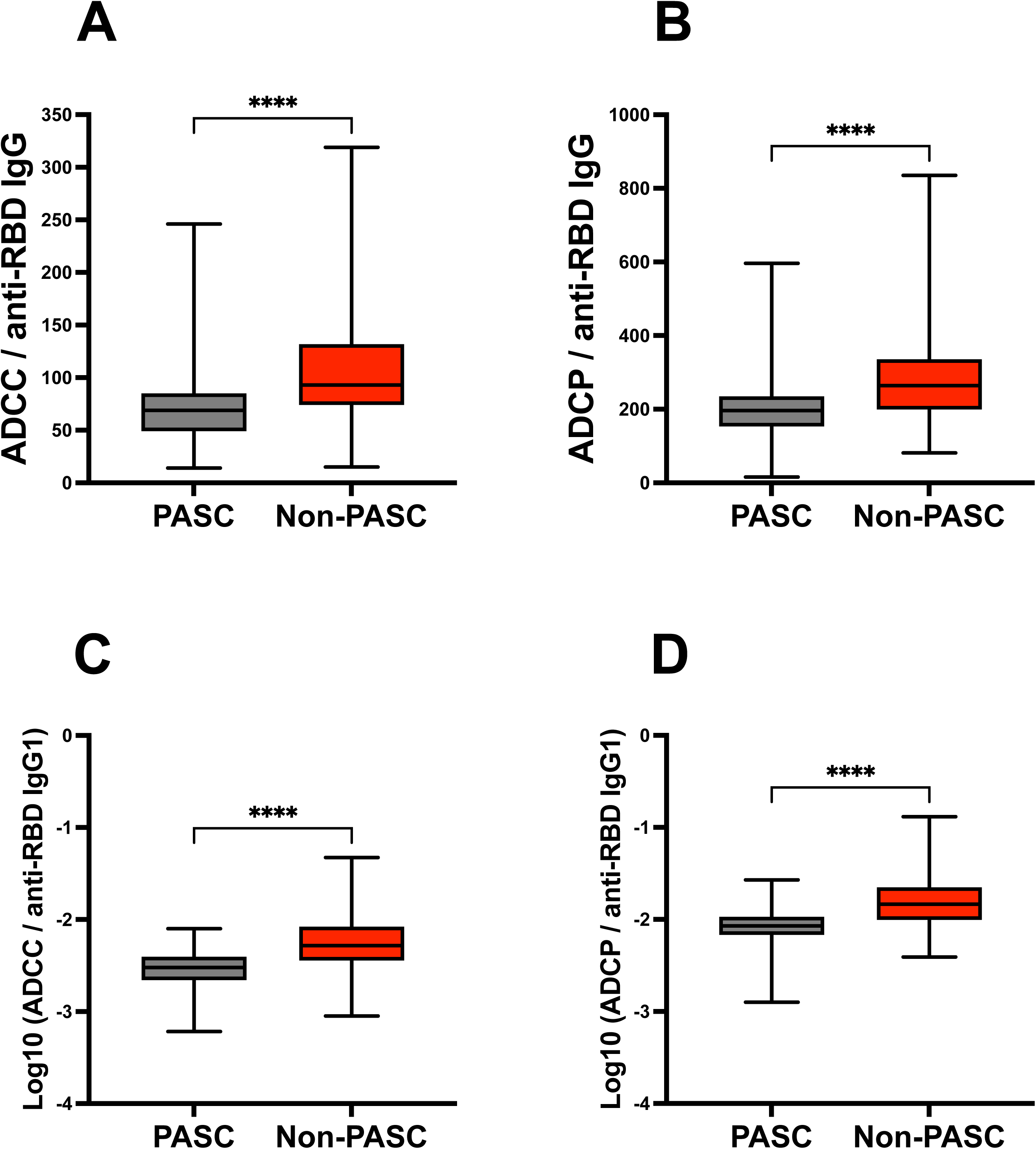
Fc-effector function of anti-RBD IgG is reduced in PASC relative to non-PASC subjects. Sera from the same RECOVER cohort (PASC, n = 99; non-PASC, n = 139) were tested in Jurkat-LuciaTM reporter-cell assays for (A) antibody-dependent cellular cytotoxicity (ADCC) and (B) antibody-dependent cellular phagocytosis (ADCP) induced by RBD-bound IgG, and for the same measures normalized to each sample’s anti-RBD IgG 1 level [(D) ADCC/IgG1, (E) ADCP/IgG1] to control for antibody abundance. Box-and-whisker plots display the median (horizontal line), interquartile range (box, 25th– 75th percentile), and full data range (whiskers). Significance levels shown are based on the p-values assessed by the Mann-Whitney U test. ****: p-value <0.0001.

**Figure 3.**
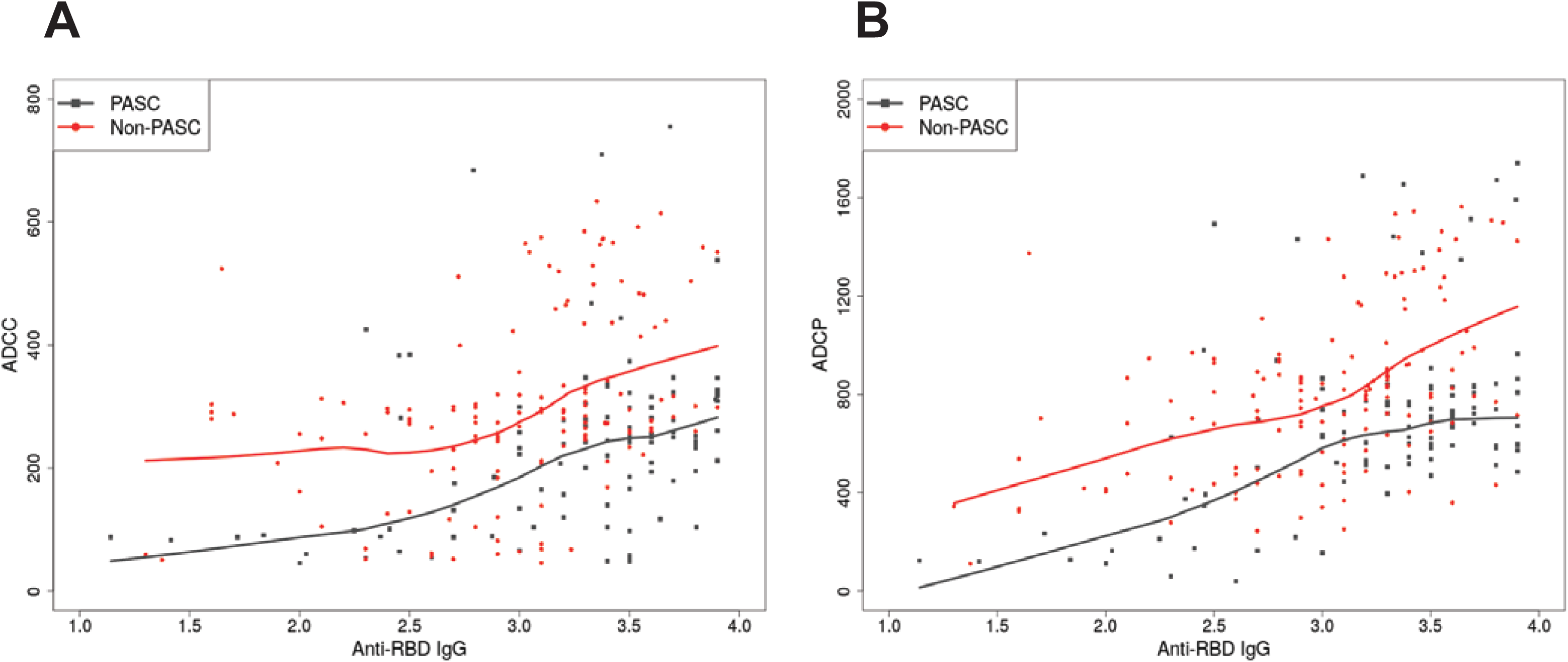
Association between anti-RBD IgG level and Fc-effector function differs between PASC and non-PASC subjects. Scatter plots show the anti-RBD IgG level versus (A) ADCC or (B) ADCP for individual serum samples from RECOVER participants with PASC (n = 99, grey squares) or without PASC (n = 139, red circles), with non-parametric (loess) trend lines for each group (grey = PASC, red =non-PASC).

### The Fc-mediated effector deficit in PASC persists after accounting for clinical covariates

Because demographic and clinical factors can influence humoral immune responses, we next asked whether the reduced FcγR-mediated antibody activity observed in PASC could be explained by differences in these variables between the two groups. Immunity-related comorbidities, which were more frequent in the PASC group, did not account for the differences in anti-RBD antibody abundance or FcγR-mediated activity within each of the groups (**Supplemental Table 1**). Multivariable linear regression analyses further examined the potential effects of age, sex, race and ethnicity, selected comorbidities, obesity, time since the most recent SARS-CoV-2 infection, time since the most recent COVID-19 vaccination, and interactions between PASC status and these time intervals. PASC status remained significantly associated with anti-RBD IgG abundance, while the relationships of anti-RBD IgG, ADCC, and ADCP with time since infection differed by PASC status (**Supplemental Table 2**); backwards and stepwise AIC optimization processes produced similar results. These findings suggest that the relationships between time since infection and both the magnitude and Fc-mediated function of the anti-RBD antibody response differ with PASC status. Importantly, after normalization of Fc-effector readouts to anti-RBD antibody abundance, the between-group functional differences remained and were not explained by the measured demographic or clinical covariates.

### Anti-RBD IgG1 is more fucosylated in PASC

Since neither antibody abundance nor measured clinical covariates explained the reduced FcγR-mediated activity in PASC, we asked whether this functional difference was associated with altered Fc glycosylation, a major structural determinant of IgG effector activity. IgG Fc effector activity is strongly regulated by N-glycosylation at Asn-297. In particular, core fucosylation reduces IgG1 affinity for FcγRIIIa (CD16a), whereas afucosylation markedly enhances FcγRIIIa binding and ADCC ^9–13^. We examined Fc glycosylation of anti-RBD IgG1 in a subset of randomly selected participants (30 PASC and 34 non-PASC). We found that the aggregate abundance of fucosylated anti-RBD IgG1 glycoforms was higher in PASC (**Figure 4A**), whereas nonfucosylated glycoforms did not differ significantly (**Figure 4B**). Thus, the reduced FcγR-mediated activity in PASC was associated with a shift toward greater Fc fucosylation.

**Figure 4.**
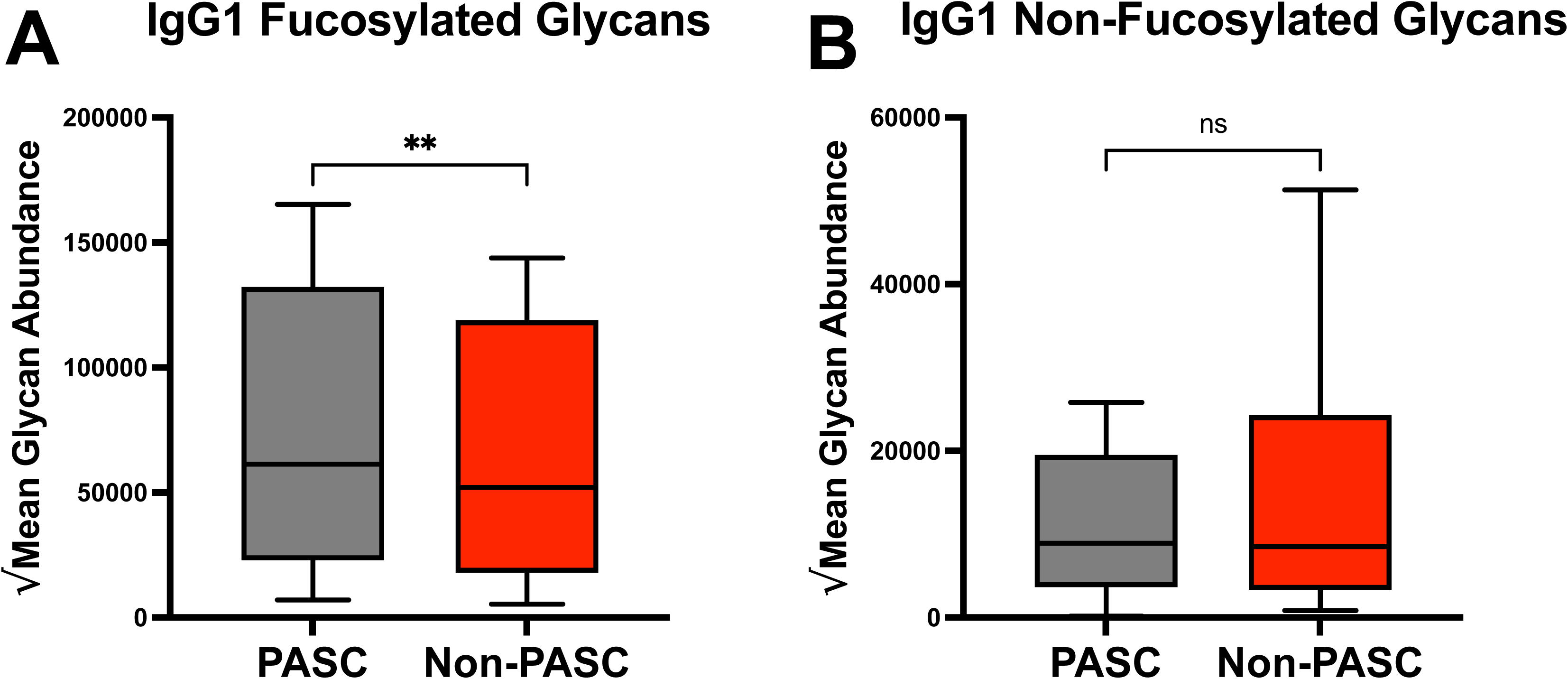
Anti-RBD IgG1 shows increased Fc fucosylation in PASC relative to non-PASC subjects. Fc N-glycan composition of anti-RBD IgG1 was determined by mass spectrometry (see Methods) in sera from PASC (n = 30) and non-PASC (n = 34) participants. Box plots summarize aggregate levels of the 10 fucosylated (A) and 11 non-fucosylated (B) glycan species detected. Boxes represent the interquartile range (25th–75th percentile); horizontal lines denote medians, and whiskers span the top and bottom quartiles of the data. For each panel, PASC and non-PASC groups were compared by a paired Wilcoxon signed-rank test, with pairing by individual glycan species (n = 10 pairs for panel A, n=11 pairs for panel B; each pair comprising the mean PASC and mean non-PASC abundance for a given glycan chain). Significance levels shown are based on the p-values from the Wilcoxon signed-rank test. **: p-value <0.0021; ns, not significant, p-value >0.05.

### Circulating FUCA2 is reduced in PASC

We next asked whether increased Fc fucosylation of anti-RBD IgG1 in PASC was accompanied by differences in circulating fucose-modifying enzymes. Core fucose is added during antibody biosynthesis by FUT8, whereas α-L-fucosidases, including FUCA1 and the secreted plasma protein FUCA2, catalyze fucose removal from glycosylated substrate^14–17^. We performed targeted quantitative mass spectrometry in a subset of participants whose samples had also been analyzed for IgG1 Fc glycosylation (24 PASC and 34 non-PASC). Serum FUCA2 concentrations were lower in PASC (**Figure 5B**), whereas FUCA1 did not differ significantly (**Figure 5A**). FUT8 was not detectable. As an additional potential modulator of FcγR activity, we also measured serum levels of soluble FcγRIIIa (CD16a), which is generated by ADAM17-mediated receptor shedding from the cell membrane and can bind circulating immune complexes, thereby limiting FcγR-mediated activity^18, 19^. Serum sCD16a levels did not differ between groups (**Figure 5D**). Thus, increased anti-RBD IgG1 fucosylation in PASC was accompanied by lower circulating levels of FUCA2, while FUCA1 and sCD16a were unchanged.

**Figure 5.**
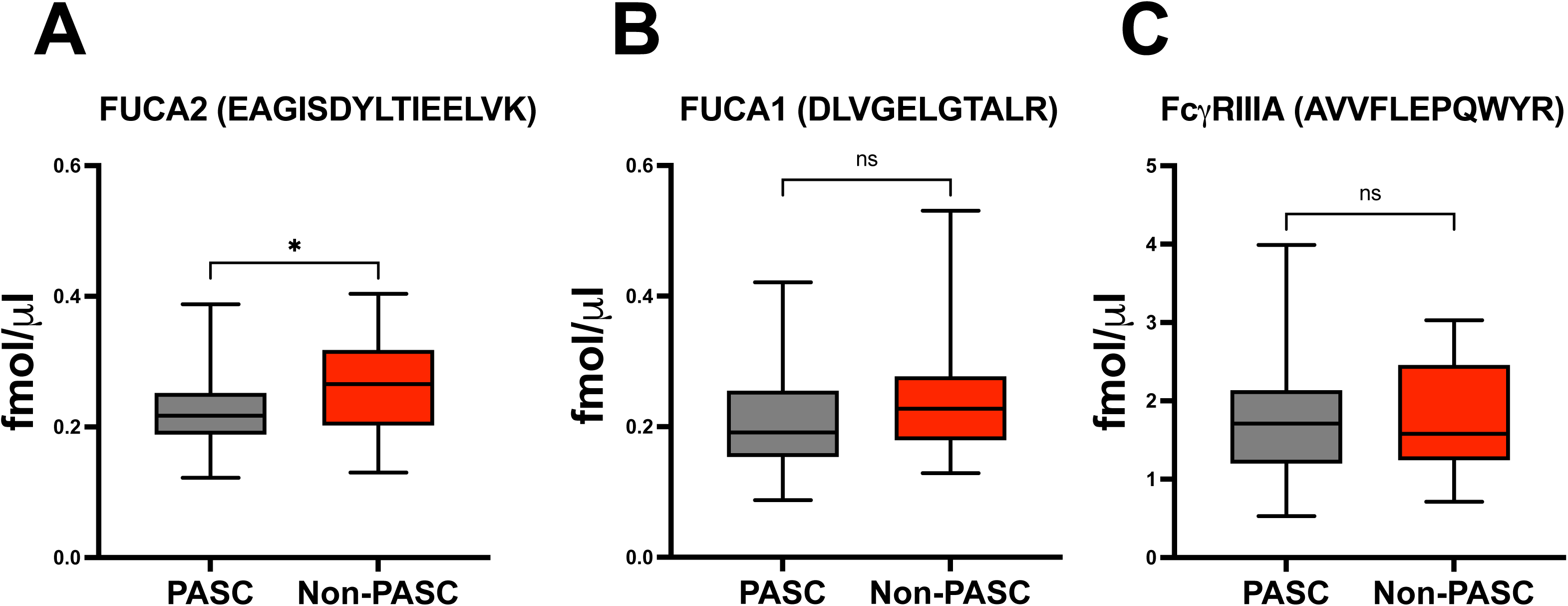
Serum concentrations of fucose-modifying enzymes and CD16a in adults with and without Post-Acute Sequelae of SARS-CoV-2 (PASC). Serum levels of (A) α-L-fucosidase 1 (FUCA1), (B) α-L-fucosidase 2 (FUCA2), and (D) the Fcγ receptor CD16a (FCGR3A) were quantified by targeted mass spectrometry (tryptic peptides DLVGELGTALR, EAGISDYLTIEELVK, and AVVFLEPQWYR, respectively; see Methods) in RECOVER participants classified as PASC (n = 24) or non-PASC (n = 34). Concentrations are expressed in femtomoles per microliter (fmol/µL). Box-and-whisker plots display the median (horizontal line), interquartile range (box, 25th–75th percentile), and full data range (whiskers). Group differences were assessed by the Mann-Whitney U test. *p < 0.05.

## Discussion

This study identifies, to our knowledge, the first PASC-associated glycoimmune signature linking impaired antiviral antibody Fc function with altered Fc glycosylation and fucose metabolism. In adult participants in the observational RECOVER study, PASC was associated with reduced FcγR-mediated antibody activity despite increased anti-RBD IgG and IgG1 abundance, increased fucosylation of anti-RBD IgG1, and reduced circulating levels of the fucose-removing enzyme FUCA2. The strong, well-established inverse relationship between IgG1 core fucosylation and FcγRIIIa activation provides a plausible basis for the reduced ADCC-associated activity observed in PASC. ADCP is also affected by Fc fucosylation, but this relationship is less straightforward and reflects the combined effects of multiple Fc glycan features, FcγR usage, and effector-cell context^20, 21^. Reduced FUCA2 further links the antibody glycosylation phenotype with altered fucose metabolism, although the mechanistic association between this enzyme and IgG Fc fucoslyation remains to be established^22^. Together, these findings reveal a qualitative alteration of the antiviral antibody response in PASC.

Reduced Fc-mediated antibody function may have important consequences for viral clearance. FcγR-dependent effector mechanisms, including ADCC and ADCP, contribute to the elimination of virus-infected cells and antigen-containing immune complexes. Attenuation of these functions could therefore impair clearance of SARS-CoV-2 antigen or infected cells, thereby contributing to antigen persistence. This possibility is particularly relevant because persistence of SARS-CoV-2 antigen or viral reservoirs is one of the leading mechanistic hypotheses for PASC. Our findings thus provide a potential antibody-based mechanism by which an apparently robust humoral response could nevertheless be functionally inadequate for efficient antigen clearance.

Several limitations should be considered. First, the cross-sectional study design precludes establishing the temporal or causal relationships among altered FcγR-mediated antibody function, increased IgG1 fucosylation, reduced circulating FUCA2, and PASC. Second, glycosylation and targeted proteomic analyses were performed in subsets of the full cohort, resulting in smaller sample sizes for these analyses. Although multivariable analyses accounted for major demographic and clinical variables, residual confounding by unmeasured factors, including acute COVID-19 severity, medication use, or PASC endotypes, cannot be excluded. In addition, FcγR-mediated function was assessed using standardized reporter cell lines rather than primary NK cells or phagocytes. Moreover, IgG function and glycosylation were evaluated only for antibodies directed against SARS-CoV-2 RBD. Whether the observed functional and glycosylation phenotypes extend to antibodies and primary effector cells directed against other antigens remains to be determined. Finally, the relationship between reduced FUCA2 and increased IgG1 fucosylation is associative; direct measurements of fucosidase activity and FUT8 expression or activity will be required to determine whether altered fucose metabolism contributes mechanistically to the antibody glycosylation phenotype in PASC.

These findings identify altered Fc-mediated antibody effector functions as potential contributors to PASC pathogenesis and provide a framework for investigating how altered Fc glycosylation and fucose metabolism affect antiviral immune function. Longitudinal and mechanistic studies will be important to define the temporal and causal relationships among these processes and to determine whether components of this glycoimmune pathway may serve as biomarkers of risk or therapeutic targets.

## Methods

### Sex as a biological variable

The study included female and male participants. Sex was included as a covariate in multivariable analyses. The study was not designed or powered to test sex-specific biological effects.

### Cohort Description

For this study, we obtained serum samples from the NIH RECOVER Initiative adult observational cohort participants --all infected and all COVID-19 vaccinated --who exhibited LC (PASC, n = 99) or did not (non-PASC, n = 139). The PASC group definition was based on history of SARS-CoV-2 infection and RECOVER Long COVID Research Index 2023 (LCRI)^23^ ≥ 12 at T2 (6 months post-infection) with only a single infection recorded and vaccinated. The non-PASC group was defined as having an LCRI < 12 at T2 (6 months post-infection). The RECOVER adult study protocol, including its rationale, objectives, and design, is described by Horwitz *et al* ^24^. The parent RECOVER-Adult protocol was approved by the IRB at NYU Grossman School of Medicine and collaborating sites, including Rutgers University (Pro2021001977). Serum specimens were received from the RECOVER cohort from Mayo Clinic, Rochester, MN.

### Enzyme-linked Immunosorbent Assay (ELISA)

Antibody binding for the IgG isotype was performed by ELISA using recombinant SARS-CoV-2 Spike RBD and N proteins as solid-phase antigens, following standard operating procedures, as described^25, 26^. For IgG subclass detection, a Luminex-based ELISA was used. Recombinant SARS-CoV-2 Spike RBD was conjugated to uniquely bar-coded carboxylated magnetic beads (MagPlex-C, Luminex Corp, Austin, TX). Bead conjugation was validated using a monoclonal antibody against SARS-CoV-2 spike (NBP2-90980PE, Novus Biologicals, Centennial, CO). Validated beads were aliquoted and stored at 4°C until use. Luminex buffer was prepared by adding 0.5 mL Tween-20 (P1379, Sigma-Aldrich, Allentown, PA) to 1 liter of Phosphate-buffered saline (PBS) (final Tween-20 concentration = 0.05%), followed by 10 g of Bovine serum albumin (BSA) to a final concentration of 1%, and 0.02% NaN3 (S227I-100, Thermo Fisher Scientific, Waltham, MA). Fifty microliters of bead slurry (RBD-coupled magnetic beads in Luminex buffer, at a final concentration of 50 beads/µL) were transferred to each well of the 96-well assay plate. The plate was blocked by incubation in 100 µL/well of 1X Luminex buffer with shaking in the dark for 30 min at room temperature. The magnetic beads were then washed twice with 100 µL/well of 1X Luminex buffer, shaken for 5 min, and placed on a magnet to separate the beads from the supernatant. Heat-inactivated serum samples (56 °C for 60 min), diluted 1:250 in 1X Luminex buffer, were added to the designated wells for each IgG subclass. After 1 h of incubation on a shaker platform at room temperature, the plates were washed twice with 100 µL/well of 1X Luminex buffer, as above. Each of four anti-human IgG subclass antibodies (50 µL each; A10630, 05-3500, 05-3600, A10651, Invitrogen, Carlsbad, CA) was added to its corresponding designated well for each sample and incubated for 30 minutes at room temperature, after which the plate was washed twice. PE-labeled anti-human IgG Fc secondary antibody (50 µL/well; 05-420-0, Invitrogen, Carlsbad, CA) was then added to each well for 30 min in the dark with shaking. Plates were then washed twice, and 150 µL of 1X Luminex buffer was added to each well, followed by overnight incubation at 4°C. Assay readout (median fluorescence intensity, MFI) was performed using a Luminex FlexMap3D^TM^ instrument (Luminex Corp, Austin, TX). Cut-off was defined as MFI values > 5 SD above the mean.

### FcγRIIIa and FcγRIIa mediated reporter cell-based assays (ADCC and ADCP surrogate assays)

Reporter cell lines Jurkat-Lucia^TM^ NFAT-CD16a for ADCC and Jurkat-Lucia^TM^ NFAT-CD32 for ADCP ((jktl-nfat-cd16, jktl-nfat-cd32) were purchased from InvivoGen, San Diego, CA. Jurkat cells naturally express a functional nuclear factor of activated T cells (NFAT), which is a transcription factor involved in the early signaling events of ADCC and ADCP. These cell lines are engineered to express high-affinity CD16a (FcγRIIIA) and CD32 (FcγRIIA). Engagement of these receptors by the antibody Fc domain initiates a signaling cascade resulting in the expression of an NFAT-inducible Lucia luciferase reporter gene. All cell lines were maintained in Roswell Park Memorial Institute (RPMI) 1640 medium (MT10104CV, Corning Inc., Corning, NY) supplemented with 10% heat-inactivated fetal bovine serum (97068-069, FBS; Seradigm, Radnor, PA), 2mM L-glutamine (IC1680149, Corning Inc., NY), and 1% penicillin/streptomycin (45000-653, Corning Inc., Corning, NY), in a controlled environment at 37°C and 5% CO_2_ in a humidified atmosphere. Blasticidin S HCl (10 µg/mL) and Zeocin (100 µg/mL) (A1113903, R25001 (Thermo Fisher Scientific, Waltham, MA) were employed to maintain the Jurkat-Lucia^TM^ cell lines. Maxi-sorp 96-well ELISA plates were coated with purified RBD (100 ng/well) overnight. Plates were washed twice with 1X PBST buffer, after which heat-inactivated test sera were added at a 1:40 dilution in 1X PBS in duplicate for 2 h at 37°C to allow immune complex formation. Controls included positive (vaccinated and infected) and negative (pre-pandemic serum) samples. Unbound antibodies (and other unwanted serum components) were removed by washing four times with 1X PBST buffer, followed by a brief PBS rinse. Subsequently, 50,000/well CD16a (for ADCC) or 50,000/well CD32 (for ADCP) Jurkat-Lucia^TM^ cells were added to the plates and incubated for 12 h in a 37°C, 5% CO_2_ incubator. Reporter luciferase production and secretion into the medium were measured by adding a coelenterazine-based Quantiluc reagent (QLCA-47-08, Invivogen, San Diego, CA) substrate solution. Luminescence values were read immediately using a BioTek Synergy Neo2 microplate reader (Agilent, Santa Clara, CA).

### Mass-spectrometric anti-RBD glycosylation detection

RBD-gp70 proteins were conjugated to NHS-activated magnetic beads in a 1 µg protein to 5 µL bead slurry ratio. Serum (50 µL) was diluted with 100 µL of binding buffer containing 150 mM Tris (pH 7.5), 170 mM NaCl, 1 mM EDTA, and 0.5% NP-40, followed by incubation with 30 µL of RBD-gp70-conjugated beads for 30 minutes at 4°C. The affinity-purified antibodies on the beads were digested with 1 µg of trypsin for 2 hours, followed by an additional 1 µg for overnight digestion. The resulting peptides were enriched and desalted using an in-house STAGE tip column^10^ packed with 2 mg of C18 beads (3 µm, Dr. Maisch GmbH, Ammerbuch, Germany), then vacuum-dried. The peptides were resuspended and analyzed on an Orbitrap Fusion mass spectrometer (Thermo Fisher Scientific, Waltham, MA) paired with an Easy-nLC 1000 nanoflow LC system (Thermo Fisher Scientific, Waltham, MA). Nano-HPLC separation used an in-house trap column (2 cm × 100 µm i.d.) and a 5 cm × 150 µm capillary column packed with 1.9 µm Reprosil-Pur Basic C18 beads (r119.b9, Dr. Maisch GmbH, Ammerbuch, Germany) under a 75-minute continuous gradient of 2–24% acetonitrile with 0.1% formic acid at 900 nL/min. Data acquisition employed data-dependent analysis (DDA) for unbiased peptide detection and targeted parallel reaction monitoring (PRM) to track 60 reported N-glucosylation sites on Asn-297 of IgG1, IgG2, and IgG4. Acquired spectra were processed in the Proteome Discoverer 2.5 interface (Thermo Fisher Scientific, Waltham, MA) using the Byonic search engine (v5.6.68, Protein Metrics, Cupertino, CA), and N-glycosylated peptide amounts were quantified by area-under-the-curve (AUC) using Skyline software (24.1.0.199, Herndon, VA). The arithmetic mean abundance was calculated separately for each glycan chain (n=10 glycan chains with fucose, n=11 glycan chains without fucose, **Table S3**) in the two groups: the PASC group (n=30) and the non-PASC group (n=34). To stabilize variance across the broad dynamic range of MS intensity data, the mean values were square-root-transformed. Consistent with established approaches for comparing paired IgG Fc glycosylation traits between clinical groups in SARS-CoV-2 cohorts^9, 27^, a paired Wilcoxon signed-rank test was conducted to compare each glycan chain between the two groups. This analysis was applied independently to both the fucosylated and non-fucosylated sets, with 10 pairs in each set.

### LC-MS acquisition and methods for serum-soluble factor detection

To obtain independent, absolute measurements of serum FUCA1, FUCA2, and FcγRIIIa, we will apply the targeted LC–MS workflow established in our preliminary studies. For each protein, at least two proteotypic peptides will be quantified using matched light (unlabeled) and stable isotope-labeled (SIL, heavy) synthetic peptides **(Table S4)**. The target peptides are selected to be unique to the human protein and absent from the rat proteome. This lets trypsin-digested rat serum serve as an analyte-free surrogate matrix for calibration standards and quality-control (QC) samples. Serum samples will be denatured, reduced, alkylated and digested with trypsin, and a fixed amount of each SIL peptide will be added prior to digestion as an internal standard. Calibration curves will be made by spiking light peptides into digested rat serum across [X–Y fmol/µL] at a constant SIL concentration. QC samples at low, mid and high concentrations will be analyzed with each batch. Peptides will be separated on a Vanquish Neo nano-UHPLC coupled to a timsTOF HT mass spectrometer operated in scheduled PRM-PASEF mode, with retention-time and ion-mobility windows set for each precursor. Data will be processed in Skyline using at least three co-eluting fragment ions per precursor. Endogenous concentrations will be calculated from light-to-heavy peak-area ratios against the matrix-matched calibration curve with appropriate weighting), then converted to protein concentration. Assay performance will be characterized by following CPTAC Tier 2 guidelines: linearity, LLOQ/ULOQ, intra- and inter-day precision (CV <20%), accuracy of QC samples (±20%), selectivity and autosampler stability.

### Statistical Analysis Methods

All relevant demographic, COVID-19 status, and resulted laboratory measures were first summarized using mean and standard deviation (SD), median with interquartile range (IQR), or as a frequency with percentage, where appropriate. These analyses were performed for the entire sample of participants as well as by PASC status. Boxplot figures were generated to visually examine the distributions of the laboratory values by PASC group. Bivariate comparisons of these measures were then separately performed to detect significant differences between PASC vs non-PASC groups using Pearson’s Chi-Square or Fisher Exact for categorical measures and Wilcoxon Rank Sum testing for continuous measures. Based on the results of the initial analyses, a series of separate multivariable linear regression models were fit with the resulted laboratory measure as outcome (Anti-RBD IgG, ADCC, ADCC/Anti-RBD IgG, ADCP, ADCP/Anti-RBD IgG, IgG1) and PASC status as primary predictor. The models were additionally adjusted for age group (18-45, 46-65, or >65 years old), sex, race/ethnicity, and relevant comorbidities (rheumatologic, autoimmune, connective tissue, or immunocompromising disease), obesity, the number of days between the most recent self-reported SARS-CoV-2 infection and sample collection, the number of days between the most recent self-reported COVID-19 vaccination and sample collection, and interactions for PASC group by days since the most recent self-reported infection and most recent self-reported vaccination.

## Data Availability

All data produced in the present study are available upon reasonable request to the authors

## Author contributions

M.L.G. supervised all aspects of the study. M.L.G., N.B., and L.C.K. conceptualized the research project and secured funding. R.U. managed the project administration. Data collection and sample processing were carried out by R.U., I.E.M., H.Z., and S.Y.J. W.H. and A.P. developed SARS-CoV-2 antigens needed for serological testing. Data analysis was performed by R.U., I.E.M., P.G., T.A., and M.L.G. Figure, table creation, and manuscript drafting were performed by R.U., P.G., T.A., and M.L.G. Recruitment of participants and data collection were performed by the RECOVER-Adults Observational Cohort Consortium. All authors reviewed the manuscript and contributed editorial feedback.

## Acknowledgments

We thank the RECOVER-Adult Observational Cohort participants and consortium investigators.

## Data availability

The authors confirm that the data supporting the findings of this study are available within the article and its supplementary information. Raw data supporting these findings can be obtained from the corresponding author upon request.

## References

1. Thomas C, Faghy MA, Owen R, Yates J, Ferraro F, Bewick T, Haggan K, Ashton REM. Lived experience of patients with Long COVID: a qualitative study in the UK. BMJ Open. 2023;13(4):e068481. Epub 20230426. doi: 10.1136/bmjopen-2022-068481. PubMed PMID: 37185640; PMCID: PMC10151237.

2. Kwissa M, Mathayan M, Salunkhe SS, Bakthavachalam V, Ye Z, Sanborn MA, Condo S, Upadhye A, Nemakal A, Wang H, Chan J, Richner JM, Basu S, Novak RM, Jacobson JR, Ganesh BB, Cerda M, Brim H, Erdmann NB, Levy BD, Marshall GD, McComsey GA, Metz TD, Okumura MJ, Peluso MJ, Walker T, Utz PJ, Krishnan JA, Prabhakar BS, Rehman J. Persistent Immune Dysregulation during Long COVID is Manifested in Antibodies Targeting Envelope and Nucleocapsid Proteins. Res Sq. 2026. Epub 20260108. doi: 10.21203/rs.3.rs-8302624/v1. PubMed PMID: 41542038; PMCID: PMC12803341.

3. Erlandson KM, Geng LN, Selvaggi CA, Thaweethai T, Chen P, Erdmann NB, Goldman JD, Henrich TJ, Hornig M, Karlson EW, Katz SD, Kim C, Cribbs SK, Laiyemo AO, Letts R, Lin JY, Marathe J, Parthasarathy S, Patterson TF, Taylor BD, Duffy ER, Haack M, Julg B, Maranga G, Hernandez C, Singer NG, Han J, Pemu P, Brim H, Ashktorab H, Charney AW, Wisnivesky J, Lin JJ, Chu HY, Go M, Singh U, Levitan EB, Goepfert PA, Nikolich JZ, Hsu H, Peluso MJ, Kelly JD, Okumura MJ, Flaherman VJ, Quigley JG, Krishnan JA, Scholand MB, Hess R, Metz TD, Costantine MM, Rouse DJ, Taylor BS, Goldberg MP, Marshall GD, Wood J, Warren D, Horwitz L, Foulkes AS, McComsey GA, Cohort RE-A. Differentiation of Prior SARS-CoV-2 Infection and Postacute Sequelae by Standard Clinical Laboratory Measurements in the RECOVER Cohort. Ann Intern Med. 2024;177(9):1209–21. Epub 20240813. doi: 10.7326/M24-0737. PubMed PMID: 39133923; PMCID: PMC11408082.

4. Proal AD, VanElzakker MB, Aleman S, Bach K, Boribong BP, Buggert M, Cherry S, Chertow DS, Davies HE, Dupont CL, Deeks SG, Eimer W, Ely EW, Fasano A, Freire M, Geng LN, Griffin DE, Henrich TJ, Iwasaki A, Izquierdo-Garcia D, Locci M, Mehandru S, Painter MM, Peluso MJ, Pretorius E, Price DA, Putrino D, Scheuermann RH, Tan GS, Tanzi RE, VanBrocklin HF, Yonker LM, Wherry EJ. SARS-CoV-2 reservoir in post-acute sequelae of COVID-19 (PASC). Nat Immunol. 2023;24(10):1616–27. Epub 20230904. doi: 10.1038/s41590-023-01601-2. PubMed PMID: 37667052.

5. Lupi L, Vitiello A, Parolin C, Calistri A, Garzino-Demo A. The Potential Role of Viral Persistence in the Post-Acute Sequelae of SARS-CoV-2 Infection (PASC). Pathogens. 2024;13(5). Epub 20240508. doi: 10.3390/pathogens13050388. PubMed PMID: 38787240; PMCID: PMC11123686.

6. Bruhns P, Jonsson F. Mouse and human FcR effector functions. Immunol Rev. 2015;268(1):25–51. doi: 10.1111/imr.12350. PubMed PMID: 26497511.

7. Cottignies-Calamarte A, Tudor D, Bomsel M. Antibody Fc-chimerism and effector functions: When IgG takes advantage of IgA. Front Immunol. 2023;14:1037033. Epub 20230202. doi: 10.3389/fimmu.2023.1037033. PubMed PMID: 36817447; PMCID: PMC9933243.

8. Hamlin RE, Pienkos SM, Chan L, Stabile MA, Pinedo K, Rao M, Grant P, Bonilla H, Holubar M, Singh U, Jacobson KB, Jagannathan P, Maldonado Y, Holmes SP, Subramanian A, Blish CA. Sex differences and immune correlates of Long COVID development, persistence, and resolution. bioRxiv. 2024. Epub 20240619. doi: 10.1101/2024.06.18.599612. PubMed PMID: 38948732; PMCID: PMC11212991.

9. Siekman SL, Pongracz T, Wang W, Nouta J, Kremsner PG, da Silva-Neto PV, Esen M, Kreidenweiss A, Held J, Trape AA, Fendel R, de Miranda Santos IKF, Wuhrer M, ImmunoCovid C. The IgG glycome of SARS-CoV-2 infected individuals reflects disease course and severity. Front Immunol. 2022;13:993354. Epub 20221018. doi: 10.3389/fimmu.2022.993354. PubMed PMID: 36389824; PMCID: PMC9641981.

10. Rappsilber J, Ishihama Y, Mann M. Stop and go extraction tips for matrix-assisted laser desorption/ionization, nanoelectrospray, and LC/MS sample pretreatment in proteomics. Anal Chem. 2003;75(3):663–70. doi: 10.1021/ac026117i. PubMed PMID: 12585499.

11. Arnold JN, Wormald MR, Sim RB, Rudd PM, Dwek RA. The impact of glycosylation on the biological function and structure of human immunoglobulins. Annu Rev Immunol. 2007;25:21–50. doi: 10.1146/annurev.immunol.25.022106.141702. PubMed PMID: 17029568.

12. Golay J, Andrea AE, Cattaneo I. Role of Fc Core Fucosylation in the Effector Function of IgG1 Antibodies. Front Immunol. 2022;13:929895. Epub 20220630. doi: 10.3389/fimmu.2022.929895. PubMed PMID: 35844552; PMCID: PMC9279668.

13. Li T, DiLillo DJ, Bournazos S, Giddens JP, Ravetch JV, Wang LX. Modulating IgG effector function by Fc glycan engineering. Proc Natl Acad Sci U S A. 2017;114(13):3485–90. Epub 20170313. doi: 10.1073/pnas.1702173114. PubMed PMID: 28289219; PMCID: PMC5380036.

14. Bastian K, Scott E, Elliott DJ, Munkley J. FUT8 Alpha-(1,6)-Fucosyltransferase in Cancer. Int J Mol Sci. 2021;22(1). Epub 20210105. doi: 10.3390/ijms22010455. PubMed PMID: 33466384; PMCID: PMC7795606.

15. Tudor L, Nedic Erjavec G, Nikolac Perkovic M, Konjevod M, Uzun S, Kozumplik O, Mimica N, Lauc G, Svob Strac D, Pivac N. The Association of the Polymorphisms in the FUT8-Related Locus with the Plasma Glycosylation in Post-Traumatic Stress Disorder. Int J Mol Sci. 2023;24(6). Epub 20230316. doi: 10.3390/ijms24065706. PubMed PMID: 36982780; PMCID: PMC10056189.

16. Prabhu SK, Li C, Zong G, Zhang R, Wang LX. Comparative studies on the substrate specificity and defucosylation activity of three alpha-l-fucosidases using synthetic fucosylated glycopeptides and glycoproteins as substrates. Bioorg Med Chem. 2021;42:116243. Epub 20210607. doi: 10.1016/j.bmc.2021.116243. PubMed PMID: 34126284; PMCID: PMC8243346.

17. Fu J, Guo Q, Feng Y, Cheng P, Wu A. Dual role of fucosidase in cancers and its clinical potential. J Cancer. 2022;13(10):3121–32. Epub 20220815. doi: 10.7150/jca.75840. PubMed PMID: 36046653; PMCID: PMC9414016.

18. Romee R, Foley B, Lenvik T, Wang Y, Zhang B, Ankarlo D, Luo X, Cooley S, Verneris M, Walcheck B, Miller J. NK cell CD16 surface expression and function is regulated by a disintegrin and metalloprotease-17 (ADAM17). Blood. 2013;121(18):3599–608. Epub 20130313. doi: 10.1182/blood-2012-04-425397. PubMed PMID: 23487023; PMCID: PMC3643761.

19. Lu F, Zhao H, Dai Y, Wang Y, Lee CH, Freeman M. Cryo-EM reveals that iRhom2 restrains ADAM17 protease activity to control the release of growth factor and inflammatory signals. Mol Cell. 2024;84(11):2152–65 e5. Epub 20240522. doi: 10.1016/j.molcel.2024.04.025. PubMed PMID: 38781971; PMCID: PMC11248996.

20. Kuhns S, Shu J, Xiang C, Guzman R, Zhang Q, Bretzlaff W, Miscalichi N, Kalenian K, Joubert M. Differential influence on antibody dependent cellular phagocytosis by different glycoforms on therapeutic Monoclonal antibodies. J Biotechnol. 2020;317:5–15. Epub 20200501. doi: 10.1016/j.jbiotec.2020.04.017. PubMed PMID: 32361021.

21. Kristic J, Lauc G. The importance of IgG glycosylation-What did we learn after analyzing over 100,000 individuals. Immunol Rev. 2024;328(1):143–70. Epub 20241004. doi: 10.1111/imr.13407. PubMed PMID: 39364834; PMCID: PMC11659926.

22. Baumges H, Jelinek S, Lange H, Markmann S, Capriotti E, Hausser JA, Ilse MB, Braulke T, Lubke T. Deciphering alpha-L-Fucosidase Activity Contribution in Human and Mouse: Tissue alpha-L-Fucosidase FUCA1 Meets Plasma alpha-L-Fucosidase FUCA2. Cells. 2025;14(17). Epub 20250830. doi: 10.3390/cells14171355. PubMed PMID: 40940767; PMCID: PMC12427801.

23. Thaweethai T, Jolley SE, Karlson EW, Levitan EB, Levy B, McComsey GA, McCorkell L, Nadkarni GN, Parthasarathy S, Singh U, Walker TA, Selvaggi CA, Shinnick DJ, Schulte CCM, Atchley-Challenner R, Alba GA, Alicic R, Altman N, Anglin K, Argueta U, Ashktorab H, Baslet G, Bassett IV, Bateman L, Bedi B, Bhattacharyya S, Bind MA, Blomkalns AL, Bonilla H, Brim H, Bush PA, Castro M, Chan J, Charney AW, Chen P, Chibnik LB, Chu HY, Clifton RG, Costantine MM, Cribbs SK, Davila Nieves SI, Deeks SG, Duven A, Emery IF, Erdmann N, Erlandson KM, Ernst KC, Farah-Abraham R, Farner CE, Feuerriegel EM, Fleurimont J, Fonseca V, Franko N, Gainer V, Gander JC, Gardner EM, Geng LN, Gibson KS, Go M, Goldman JD, Grebe H, Greenway FL, Habli M, Hafner J, Han JE, Hanson KA, Heath J, Hernandez C, Hess R, Hodder SL, Hoffman MK, Hoover SE, Huang B, Hughes BL, Jagannathan P, John J, Jordan MR, Katz SD, Kaufman ES, Kelly JD, Kelly SW, Kemp MM, Kirwan JP, Klein JD, Knox KS, Krishnan JA, Kumar A, Laiyemo AO, Lambert AA, Lanca M, Lee-Iannotti JK, Logarbo BP, Longo MT, Luciano CA, Lutrick K, Maley JH, Mallett G, Marathe JG, Marconi V, Marshall GD, Martin CF, Matusov Y, Mehari A, Mendez-Figueroa H, Mermelstein R, Metz TD, Morse R, Mosier J, Mouchati C, Mullington J, Murphy SN, Neuman RB, Nikolich JZ, Ofotokun I, Ojemakinde E, Palatnik A, Palomares K, Parimon T, Parry S, Patterson JE, Patterson TF, Patzer RE, Peluso MJ, Pemu P, Pettker CM, Plunkett BA, Pogreba-Brown K, Poppas A, Quigley JG, Reddy U, Reece R, Reeder H, Reeves WB, Reiman EM, Rischard F, Rosand J, Rouse DJ, Ruff A, Saade G, Sandoval GJ, Santana JL, Schlater SM, Sciurba FC, Shepherd F, Sherif ZA, Simhan H, Singer NG, Skupski DW, Sowles A, Sparks JA, Sukhera FI, Taylor BS, Teunis L, Thomas RJ, Thorp JM, Thuluvath P, Ticotsky A, Tita AT, Tuttle KR, Urdaneta AE, Valdivieso D, VanWagoner TM, Vasey A, Verduzco-Gutierrez M, Wallace ZS, Ward HD, Warren DE, Weiner SJ, Welch S, Whiteheart SW, Wiley Z, Wisnivesky JP, Yee LM, Zisis S, Horwitz LI, Foulkes AS, Consortium R. Development of a Definition of Postacute Sequelae of SARS-CoV-2 Infection. JAMA. 2023;329(22):1934–46. doi: 10.1001/jama.2023.8823. PubMed PMID: 37278994; PMCID: PMC10214179.

24. Horwitz LI, Thaweethai T, Brosnahan SB, Cicek MS, Fitzgerald ML, Goldman JD, Hess R, Hodder SL, Jacoby VL, Jordan MR, Krishnan JA, Laiyemo AO, Metz TD, Nichols L, Patzer RE, Sekar A, Singer NG, Stiles LE, Taylor BS, Ahmed S, Algren HA, Anglin K, Aponte-Soto L, Ashktorab H, Bassett IV, Bedi B, Bhadelia N, Bime C, Bind MC, Black LJ, Blomkalns AL, Brim H, Castro M, Chan J, Charney AW, Chen BK, Chen LQ, Chen P, Chestek D, Chibnik LB, Chow DC, Chu HY, Clifton RG, Collins S, Costantine MM, Cribbs SK, Deeks SG, Dickinson JD, Donohue SE, Durstenfeld MS, Emery IF, Erlandson KM, Facelli JC, Farah-Abraham R, Finn AV, Fischer MS, Flaherman VJ, Fleurimont J, Fonseca V, Gallagher EJ, Gander JC, Gennaro ML, Gibson KS, Go M, Goodman SN, Granger JP, Greenway FL, Hafner JW, Han JE, Harkins MS, Hauser KSP, Heath JR, Hernandez CR, Ho O, Hoffman MK, Hoover SE, Horowitz CR, Hsu H, Hsue PY, Hughes BL, Jagannathan P, James JA, John J, Jolley S, Judd SE, Juskowich JJ, Kanjilal DG, Karlson EW, Katz SD, Kelly JD, Kelly SW, Kim AY, Kirwan JP, Knox KS, Kumar A, Lamendola-Essel MF, Lanca M, Lee-Lannotti JK, Lefebvre RC, Levy BD, Lin JY, Logarbo BP, Jr., Logue JK, Longo MT, Luciano CA, Lutrick K, Malakooti SK, Mallett G, Maranga G, Marathe JG, Marconi VC, Marshall GD, Martin CF, Martin JN, May HT, McComsey GA, McDonald D, Mendez-Figueroa H, Miele L, Mittleman MA, Mohandas S, Mouchati C, Mullington JM, Nadkarni GN, Nahin ER, Neuman RB, Newman LT, Nguyen A, Nikolich JZ, Ofotokun I, Ogbogu PU, Palatnik A, Palomares KTS, Parimon T, Parry S, Parthasarathy S, Patterson TF, Pearman A, Peluso MJ, Pemu P, Pettker CM, Plunkett BA, Pogreba-Brown K, Poppas A, Porterfield JZ, Quigley JG, Quinn DK, Raissy H, Rebello CJ, Reddy UM, Reece R, Reeder HT, Rischard FP, Rosas JM, Rosen CJ, Rouphael NG, Rouse DJ, Ruff AM, Saint Jean C, Sandoval GJ, Santana JL, Schlater SM, Sciurba FC, Selvaggi C, Seshadri S, Sesso HD, Shah DP, Shemesh E, Sherif ZA, Shinnick DJ, Simhan HN, Singh U, Sowles A, Subbian V, Sun J, Suthar MS, Teunis LJ, Thorp JM, Jr., Ticotsky A, Tita ATN, Tragus R, Tuttle KR, Urdaneta AE, Utz PJ, VanWagoner TM, Vasey A, Vernon SD, Vidal C, Walker T, Ward HD, Warren DE, Weeks RM, Weiner SJ, Weyer JC, Wheeler JL, Whiteheart SW, Wiley Z, Williams NJ, Wisnivesky JP, Wood JC, Yee LM, Young NM, Zisis SN, Foulkes AS. Researching COVID to Enhance Recovery (RECOVER) adult study protocol: Rationale, objectives, and design. PLoS One. 2023;18(6):e0286297. Epub 20230623. doi: 10.1371/journal.pone.0286297. PubMed PMID: 37352211; PMCID: PMC10289397.

25. Bruiners N, Ukey R, Konvinse KC, Harris M, Kalaycioglu M, Yang JH, Yang E, Ganapathi U, Honnen W, Andrews T, Richlin B, Suarez C, Gaur S, Kalyoussef S, Ricciardi E, Hasan UN, Cuddy W, Singh AR, Wahezi D, Rothschild E, Brady PW, Lakhani SA, Bukulmez H, Kaelber DC, Kimura Y, Pinter A, Napoli S, Moroso-Fela S, Kleinman LC, Horton DB, Utz PJ, Gennaro ML. Antibody repertoire associated with clinically diverse presentations of pediatric SARS-CoV-2 infection. Sci Rep. 2026. Epub 20260612. doi: 10.1038/s41598-026-56351-6. PubMed PMID: 42286032.

26. Datta P, Ukey R, Bruiners N, Honnen W, Carayannopoulos MO, Reichman C, Choudhary A, Onyuka A, Handler D, Guerrini V, Mishra PK, Dewald HK, Lardizabal A, Lederer L, Leiser AL, Hussain S, Jagpal SK, Radbel J, Bhowmick T, Horton DB, Barrett ES, Xie YL, Fitzgerald-Bocarsly P, Weiss SH, Woortman M, Parmar H, Roy J, Dominguez-Bello MG, Blaser MJ, Carson JL, Panettieri RA, Jr., Libutti SK, Raymond HF, Pinter A, Gennaro ML. Highly versatile antibody binding assay for the detection of SARS-CoV-2 infection and vaccination. J Immunol Methods. 2021;499:113165. Epub 20211009. doi: 10.1016/j.jim.2021.113165. PubMed PMID: 34634317; PMCID: PMC8500840.

27. Pongracz T, Nouta J, Wang W, van Meijgaarden KE, Linty F, Vidarsson G, Joosten SA, Ottenhoff THM, Hokke CH, de Vries JJC, Arbous SM, Roukens AHE, Wuhrer M, Beat C, groups C-. Immunoglobulin G1 Fc glycosylation as an early hallmark of severe COVID-19. EBioMedicine. 2022;78:103957. Epub 20220322. doi: 10.1016/j.ebiom.2022.103957. PubMed PMID: 35334306; PMCID: PMC8938159.

