## Supplementary Information for "Antiviral Antibody Fc Dysfunction Is Linked to Increased IgG1 Fucosylation in Long COVID"

#### Supplementary Figure legends

**Figure S1**. Association between anti-RBD IgG1 level and Fc-effector function differs between PASC and non-PASC subjects. Scatter plots show the anti-RBD IgG1 level versus (A) ADCC or (B) ADCP for individual serum samples from RECOVER participants with PASC (n = 99, grey squares) or without PASC (n = 139, red circles), with non-parametric (loess) smoothed trend lines for each group (grey = PASC, red =non-PASC).

#### Supplementary Tables

**Supplementary Table 1.** Laboratory outcome measures by PASC status and immunocompromising, rheumatologic/autoimmune comorbidity status.

**Supplementary Table 2.** Multivariable linear regression models of laboratory outcomes in PASC versus non-PASC participants.

**Supplementary Table 3.** List of glycan chains by the presence of fucose.

**Supplementary Table 4.** List of synthetic peptides used for targeted mass spectrometry.

### Figure S1

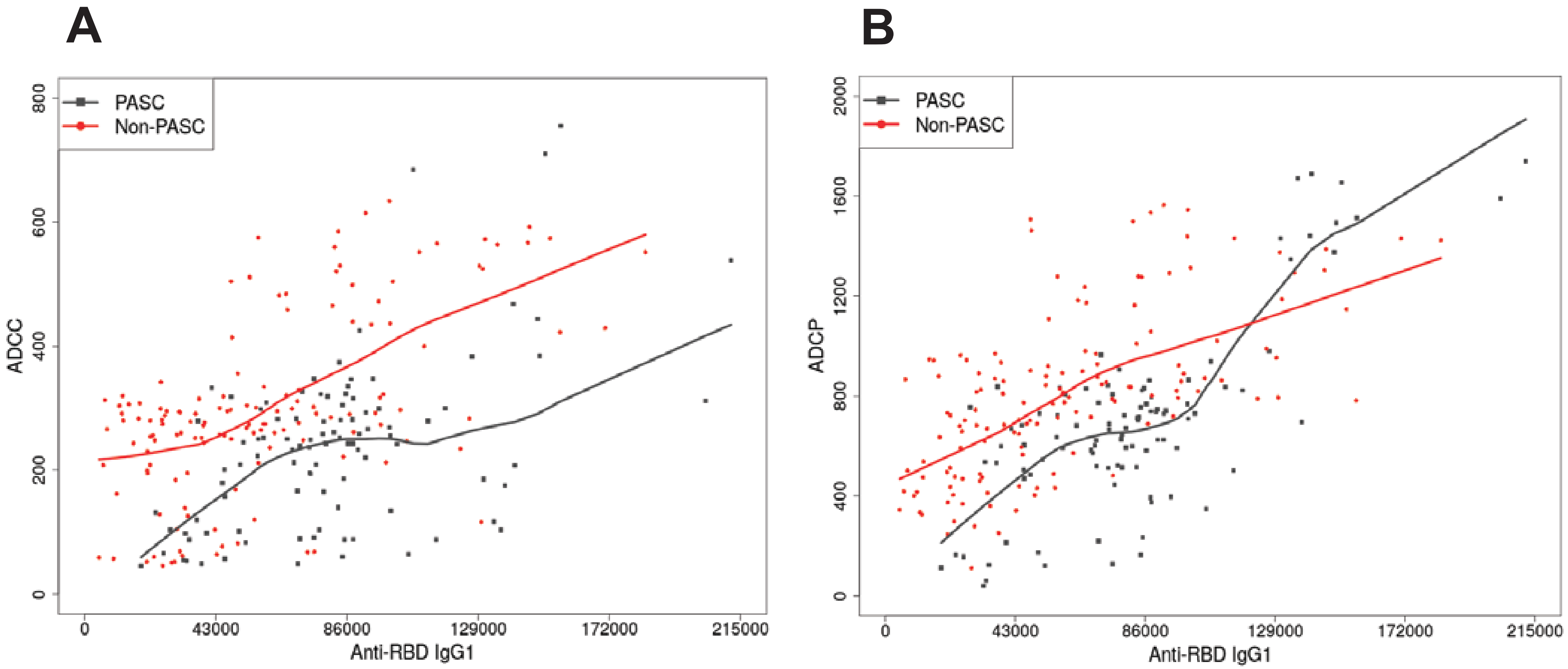
A
B
PASC
Non-PASC
ADCC
ADCP
0
200
400
600
800
1200
1600
2000
43000
86000
129000
172000
215000
Anti-RBD IgG1

**Table S1. Laboratory outcome measures by PASC status and immunocompromising, rheumatologic/autoimmune comorbidity status.**

| | PASC | | | Non-PASC | | |
|---|---|---|---|---|---|---|
| | No Immunocompromising, Rheumatologic, or Auto-Immune Comorbidity | Any Immunocompromising, Rheumatologic, or Auto-Immune Comorbidity | P-Value* | No Immunocompromising, Rheumatologic, or Auto-Immune Comorbidity | Any Immunocompromising, Rheumatologic, or Auto-Immune Comorbidity | P-Value* |
| | n = 57 | n = 42 | | n = 120 | n = 19 | |
| **Anti-RBD IgG** | | | p = 0.83 | | | p = 0.22 |
| Range | 1.1 - 3.9 | 1.4 - 3.9 | | 1.3 - 3.8 | 1.4 - 3.9 | |
| Mean (SD) | 3.2 (0.6) | 3.2 (0.6) | | 3.0 (0.5) | 3.1 (0.6) | |
| Median [Q1, Q3] | 3.4 [3.0, 3.6] | 3.4 [3.0, 3.6] | | 3.1 [2.7, 3.3] | 3.1 [2.8, 3.6] | |
| **ADCC** | | | p = 0.89 | | | p = 0.62 |
| Range | 46.0 - 755.5 | 49.0 - 685.0 | | 46.0 - 634.0 | 51.0 - 614.5 | |
| Mean (SD) | 243.0 (141.8) | 232.8 (126.9) | | 302.4 (136.2) | 290.0 (189.4) | |
| Median [Q1, Q3] | 243.0 [134.5, 305.0] | 243.5 [134.0, 298.8] | | 284.0 [247.5, 323.8] | 260.0 [115.0, 414.2] | |
| **ADCC/Anti-RBD IgG** | | | p = 0.69 | | | p = 0.15 |
| Range | 14.0 - 210.5 | 14.4 - 245.5 | | 14.8 - 318.7 | 21.3 - 168.7 | |
| Mean (SD) | 73.7 (39.0) | 72.6 (43.7) | | 103.0 (47.1) | 88.0 (51.1) | |
| Median [Q1, Q3] | 71.1 [48.4, 86.9] | 68.3 [49.5, 83.2] | | 95.1 [76.3, 129.0] | 76.7 [44.8, 130.0] | |
| **ADCP** | | | p = 0.82 | | | p = 0.66 |
| Range | 41.0 - 1739.5 | 121.0 - 1670.5 | | 245.0 - 1545.5 | 111.5 - 1563.5 | |
| Mean (SD) | 691.3 (370.8) | 710.6 (376.9) | | 809.1 (310.4) | 802.6 (424.9) | |
| Median [Q1, Q3] | 696.0 [513.0, 823.0] | 635.8 [525.5, 797.0] | | 805.0 [586.8, 948.2] | 722.0 [472.0, 1091.2] | |
| **ADCP/Anti-RBD IgG** | | | p = 0.89 | | | p = 0.25 |
| Range | 15.8 - 529.7 | 52.0 - 596.4 | | 81.3 - 835.4 | 81.1 - 429.2 | |
| Mean (SD) | 207.9 (103.6) | 216.8 (114.9) | | 273.9 (102.0) | 245.8 (106.1) | |
| Median [Q1, Q3] | 196.3 [158.6, 228.8] | 196.7 [151.8, 235.2] | | 264.8 [202.1, 335.2] | 229.2 [168.4, 340.5] | |

| | | | | | | |
|---|---|---|---|---|---|---|
| **IgG1** | | | p = 0.58 | | | p = 0.19 |
| Range | 18528.0 - 211939.0 | 23225.0 - 203545.0 | | 4750.0 - 155808.0 | 21424.0 - 183826.0 | |
| Mean (SD) | 80679.4 (35901.4) | 83561.2 (36087.5) | | 57259.5 (35512.4) | 76572.3 (52201.6) | |
| Median [Q1, Q3] | 78342.0 [56909.0, 88093.0] | 81351.0 [60608.5, 95931.5] | | 53149.5 [29120.8, 81414.5] | 50182.0 [32688.0, 105018.0] | |
| **IgG2** | | | p = 0.82 | | | p = 0.26 |
| Range | 3176.0 - 243715.0 | 3893.0 - 396920.0 | | 1668.0 - 780737.0 | 5383.0 - 220828.0 | |
| Mean (SD) | 56805.6 (65467.6) | 59366.0 (74692.7) | | 56713.2 (94972.4) | 55789.0 (53273.5) | |
| Median [Q1, Q3] | 22390.0 [9897.0, 78173.0] | 27870.5 [11976.5, 88145.0] | | 17080.0 [8251.2, 72020.0] | 53353.0 [10643.0, 80935.0] | |
| **IgG3** | | | p = 0.83 | | | p = 0.048 |
| Range | 2969.0 - 53980.0 | 2887.0 - 146385.0 | | 1688.0 - 85197.0 | 2127.0 - 122837.0 | |
| Mean (SD) | 14405.0 (11677.0) | 18263.4 (24890.2) | | 8255.6 (9627.3) | 16524.3 (27403.2) | |
| Median [Q1, Q3] | 10971.0 [5511.0, 18130.0] | 10352.0 [5295.2, 18706.8] | | 5085.5 [3311.5, 9794.2] | 7175.0 [4398.0, 15875.5] | |
| **IgG4** | | | p = 0.92 | | | p = 0.28 |
| Range | 1149.0 - 979392.0 | 1408.0 - 1002070.0 | | 1779.0 - 1884930.0 | 1660.0 - 1624324.0 | |
| Mean (SD) | 309951.8 (287448.6) | 319859.6 (307010.4) | | 335589.5 (441571.5) | 409673.8 (424935.2) | |
| Median [Q1, Q3] | 281776.0 [32176.0, 518858.0] | 263363.6 [7990.8, 557142.8] | | 231346.5 [22835.5, 376994.2] | 282810.0 [143249.5, 416792.0] | |

*All p-values were resulted using a Wilcoxon Rank Sum Test

#### Table S2. Multivariable linear regression models of laboratory outcomes in PASC versus non-PASC participants.

| | PASC vs Non-PASC | | | | | |
|---|---|---|---|---|---|---|
| | **Model 1** | **Model 2** | **Model 3** | **Model 4** | **Model 5** | **Model 6** |
| | **Outcome = Anti-RBD IgG** | **Outcome = ADCC** | **Outcome = ADCC/Anti-RBD IgG** | **Outcome = ADCP** | **Outcome = ADCP/Anti-RBD IgG** | **Outcome = IgG1** |
| **Predictors** | **Beta Estimate (SE)** | | | | | |
| (Intercept) | 2.866 (0.147) | 212.597 (36.246) | 78.263 (11.596) | 634.992 (91.003) | 238.078 (27.792) | 41071.030 (9756.090) |
| Group = PASC | **0.578 (0.183)**** | -10.520 (45.042) | -16.822 (14.410) | 30.134 (113.088) | -35.705 (34.537) | 20837.200 (12123.770) |
| Age Category = 46-65 | 0.076 (0.084) | 1.762 (20.717) | -2.693 (6.628) | 36.321 (52.013) | 3.596 (15.885) | 8171.780 (5576.170) |
| Age Category = >65 | 0.154 (0.108) | -8.086 (26.611) | -9.587 (8.513) | 122.092 (66.813) | 16.352 (20.404) | 7445.590 (7162.760) |
| Sex Category = Male | -0.013 (0.080) | -7.114 (19.783) | -2.178 (6.329) | -61.601 (49.67) | -22.149 (15.169) | -1076.800 (5324.980) |
| Race Group = Hispanic | 0.205 (0.141) | 15.240 (34.699) | -0.834 (11.101) | 24.13 (87.118) | -5.096 (26.606) | -11452.440 (9339.650) |
| Race Group = Mixed race/Other/Missing | 0.022 (0.165) | -30.509 (40.595) | -10.797 (12.987) | -37.051 (101.922) | -10.900 (31.126) | 12622.750 (10926.650) |
| Race Group = Non-Hispanic Asian | 0.108 (0.235) | 34.073 (57.942) | 8.952 (18.536) | 214.783 (145.474) | 67.078 (44.427) | 24120.310 (15595.800) |
| Race Group = Non-Hispanic Black | -0.097 (0.112) | 52.158 (27.646) | 15.665 (8.844) | 57.842 (69.41) | 21.810 (21.198) | 2375.120 (7441.240) |
| Has 1+ Immuno/Rheum/Auto-immune Comorbidities = Yes | 0.049 (0.091) | 8.065 (22.415) | -0.283 (7.171) | 11.833 (56.277) | -4.441 (17.187) | 9398.430 (6033.250) |
| Days since most recent COVID infection | 0.004 (0.003) | **1.570 (0.622)**** | 0.284 (0.199) | **3.855 (1.561)**** | 0.724 (0.477) | 303.010 (167.310) |
| Days since most recent COVID vaccination | 0.000 (0.000) | 0.205 (0.118) | **0.088 (0.038)**** | 0.179 (0.296) | 0.063 (0.090) | 15.860 (31.750) |
| Group = PASC * Days since most recent COVID infection | **-0.008 (0.003)**** | **-2.182 (0.740)**** | -0.400 (0.237) | **-3.962 (1.857)**** | -0.536 (0.567) | -156.600 (199.060) |
| Group = PASC * Days since most recent COVID vaccination | 0.000 (0.001) | 0.004 (0.158) | -0.014 (0.051) | -0.153 (0.398) | -0.046 (0.121) | 17.540 (42.630) |

**p-value < 0.05, categorical predictor reference levels for all models were: Group = Non-PASC, Age Category = 18-45 years old, Sex Category = Female, Race Group = Non-Hispanic White, and Has 1+ Immuno/Rheum/Auto-immune Comorbidities = No

**Table S3**. **List of glycan chains by the presence of fucose.**

| Fucosylated Glycans | Non-fucosylated Glycans |
|---|---|
| IgG1_GoF | IgG1_G0 |
| IgG1_GoFN | IgG1_GON |
| IgG1_G1F | IgG1_G1 |
| IgG1_G1FN | IgG1_G1N |
| IgG1_G1FNS | IgG1_G1NS |
| IgG1_G1FS | IgG1_G1S |
| IgG1_G2F | IgG1_G2 |
| IgG1_G2FN | IgG1_G2N |
| IgG1_G2FNS | IgG1_G2N |
| IgG1_G2FS | IgG1_G2NS |
| | IgG1_G2S |

**Table S4**. **List of synthetic peptides used for targeted mass spectrometry.**

| Peptide Name | Sequence<br>() means labeled, [C]: modified | Length |
|---|---|---|
| FUT8-1 | YPTYPEAE(K) | 9 |
| FUT8-1 | VHGDPAVWWVSQFV(K) | 15 |
| FUCA1-1 | ITMLGIQGDL(K) | 11 |
| FUCA1-2 | DGLIVPIFQE(R) | 11 |
| FUCA2-1 | FDPTWESLDA(R) | 11 |
| FUCA2-2 | EAGISDYLTIEELV(K) | 15 |
| FUCA2-3 | VNGEAIYETHTW(R) | 13 |
| FCGR3A-1 | AVVFLEPQWY(R) | 11 |
| FCGR3A-2 | DSGSYF[C](R) (C: Carbamidomethyl) | 8 |
| FUT6 | YYQSLQAHL(K) | 10 |
